# Vaccine effectiveness of Ad26.COV2.S against symptomatic COVID-19 and clinical outcomes in Brazil: a test-negative study design

**DOI:** 10.1101/2021.10.15.21265006

**Authors:** Otavio T Ranzani, Rogério dos Santos Leite, Larissa Domingues Castilho, Crhistinne Cavalheiro Maymone Gonçalves, Geraldo Resende, Rosana Leite de Melo, Julio Croda

**Author notes:** Correspondence to: Prof Julio Croda, Universidade Federal de Mato Grosso do Sul and Fundação Oswaldo Cruz.

## Abstract

We used a test-negative design to estimate the vaccine effectiveness of Ad26.COV2.S (Janssen) against symptomatic COVID-19 and clinical outcomes in Mato-Grosso do Sul, Brazil. We analyzed 11,817 RT-PCR tests. The mean age was 37 (SD=17) years, 2,308 (20%) of individuals more or equal than 50 years and almost two-thirds of the population was Brown/Pardo. Adjusted effectiveness against symptomatic COVID-19 after 28 days of the single dose was 50.9% (95% CI, 35.5-63.0). Adjusted effectiveness against clinical outcomes was 72.9% (95% CI, 35.1-91.1) for hospitalization, 92.5% (95% CI, 54.9-99.6) for ICU admission, 88.7% (95% CI, 17.9-99.5) for mechanical ventilation and 90.5% (95% CI, 31.5-99.6) for death. Despite lacking precision on some estimates, a single dose of Ad26.COV2.S vaccine continues to protect specially for severe forms of COVID-19 in the context of new variants.

## Introduction

Ad26.COV2.S (Janssen, Johnson & Johnson) is an adenoviral vaccine against COVID-19, but unlike all other vaccine platforms, it is administered in a single dose.^1^ There is limited real-world effectiveness data for Ad26.COV2.S,^2,3^ and a need for more studies for this vaccine especially in the context of Gamma and Delta variants. We used a test-negative design to estimate the vaccine effectiveness of Ad26.COV2.S against symptomatic COVID19 and clinical outcomes in Mato-Grosso do Sul, a central-west state in Brazil with borders to Bolivia and Paraguay.

## Methods

We analyzed data from 25-06-2021 to 30-09-2021 in Mato-Grosso do Sul state (∼2.8 million inhabitants), Brazil. In Mato-Grosso do Sul, Ad26.COV2.S started to be distributed on 25-06-2021 by the national campaign program. Additionally, there was a massive campaign vaccination with Ad26.COV2.S in 13 municipalities that comprises the international borders. During June, 2021, Mato-Grosso do Sul faced a huge COVID-19 Gamma outbreak.^4^

We followed the same methodology as previously described regarding inclusion/exclusion criteria.^5,6^ Briefly, we included RT-PCR from adults residing in Mato-Grosso do Sul, collected within 10 days of symptoms onset, and excluded previous infected individuals. Our primary outcome is vaccine effectiveness after 28 days of the single dose against symptomatic COVID-19. We defined a symptomatic case as presenting at least two acute respiratory symptoms, as defined in the efficacy trial of Ad26.COV2.S.^1^ Additionally, we analyzed effectiveness against hospital and ICU admissions, use of invasive mechanical ventilation and death.^6^

We estimated vaccine effectiveness as 1-OR from logistic regression models. We adjusted the models for age, sex, diabetes mellitus, renal, hepatic, neurologic, hematologic, cardiovascular and respiratory chronic diseases, obesity, immunosuppressed status and time (day of year). Age and day of year were modelled with a restricted cubic spline. We used the period 0-13 days after the single dose as a bias-indicator.^5,6^ All analyses were conducted in R statistical software version 4.0.3 (R Project for Statistical Computing). The study was approved by the Ethical Committee for Research of Federal University of Mato-Grosso do Sul (CAAE: 43289221.5.0000.0021).

## Results

We analyzed 11,817 RT-PCR tests. The mean age was 37±17 years, 2,308 (20%) of individuals ≥50 years and almost two-thirds of the population was Brown/Pardo. The prevalence of chronic comorbidities was low, being most frequent Diabetes mellitus (2.5%)(**Table 1**).

**Table 1.** Characteristics of the analyzed population.

|  | Category | Overall | Test-negative | Test-positive |
| --- | --- | --- | --- | --- |
|  |  | 11817 | 7589 | 4228 |
| **Age** | Mean (SD), years | 37.7 (17) | 37.0 (17) | 38.9 (17) |
| **Sex, n (%)** | Female | 6181 (52.3) | 4101 (54.0) | 2080 (49.2) |
|  | Male | 5636 (47.7) | 3488 (46.0) | 2148 (50.8) |
| **Self-reported race, n (%)\*** | White/Branca | 3440 (35.6) | 2192 (35.5) | 1248 (35.7) |
|  | Brown/Pardo | 5631 (58.2) | 3618 (58.5) | 2013 (57.6) |
|  | Black/Preta | 323 (3.3) | 201 (3.3) | 122 (3.5) |
|  | Yellow/Amarela | 249 (2.6) | 151 (2.4) | 98 (2.8) |
|  | Indigenous/Indígena | 32 (0.3) | 21 (0.3) | 11 (0.3) |
| **Chronic comorbidities, n (%)** |  |  |  |  |
| Diabetes mellitus | Yes | 298 (2.5) | 166 (2.2) | 132 (3.1) |
| Renal | Yes | 36 (0.3) | 26 (0.3) | 10 (0.2) |
| Cardiovascular | Yes | 485 (4.1) | 277 (3.7) | 208 (4.9) |
| Respiratory | Yes | 174 (1.5) | 122 (1.6) | 52 (1.2) |
| Imunodepressed status | Yes | 75 (0.6) | 58 (0.8) | 17 (0.4) |
| Obesity | Yes | 135 (1.1) | 59 (0.8) | 76 (1.8) |
| Liver | Yes | 7 (0.1) | 2 (0.0) | 5 (0.1) |
| Neurologic | Yes | 24 (0.2) | 10 (0.1) | 14 (0.3) |
| Hematologic | Yes | 4 (<0.1) | 2 (<0.1) | 2 (<0.1) |
| **Place of residence, n (%)** | Other | 4994 (42.3) | 3207 (42.3) | 1787 (42.3) |
|  | Capital | 6308 (53.4) | 4061 (53.5) | 2247 (53.1) |
|  | Massive | 515 (4.4) | 321 (4.2) | 194 (4.6) |
| **From symptoms onset to RT-PCR** | Median [p25-75], Days | 4 [3, 5] | 4 [3, 5] | 4 [3, 5] |
| **Hospital admission, n (%)** | Yes | 827/11782 (7.0) | 349/7570 (4.6) | 478/4212 (11.4) |
| **ICU admission, n (%)** | Yes | 238/11700 (2.0) | 82/7520 (1.1) | 156/4180 (3.7) |
| **Use of invasive mechanical ventilation, n (%)** | Yes | 166/11694 (1.4) | 58/7522 (0.8) | 108/4172 (2.6) |
| **Death, n (%)** | Yes | 214/11586 (1.9) | 75/7444 (1.0) | 139/4142 (3.4) |
| **From symptoms onset to hospitalization** | Median [p25-75], Days | 5 [2, 7] | 3 [1, 6] | 6 [3, 8] |
| **From symptoms onset to death** | Median [p25-75], Days | 16 [7, 26] | 10 [3, 19] | 18 [11, 27] |
| **Vaccination** |  |  |  |  |
| Vaccine status, n (%) | Unvaccinated | 10655 (90.2) | 6829 (90.0) | 3826 (90.5) |
|  | 0-13 days after single dose | 430 (3.6) | 214 (2.8) | 216 (5.1) |
|  | 14-27 days after single dose | 373 (3.2) | 258 (3.4) | 115 (2.7) |
|  | ≥28 days after single dose | 359 (3.0) | 288 (3.8) | 71 (1.7) |
| From vaccination to RT-PCR | Median [p25-75], Days | 18 [11, 31] | 23 [13, 35] | 13 [9, 22] |

\* There are 2,142 missing values for self-reported race, equally distributed between test-negative and test-positive

Adjusted effectiveness against symptomatic COVID-19 after 28 days of the single dose was 50.9% (95% CI, 35.5-63.0). Adjusted effectiveness against clinical outcomes was 72.9% (95% CI, 35.1-91.1) for hospitalization, 92.5% (95% CI, 54.9-99.6) for ICU admission, 88.7% (95% CI, 17.9-99.5) for mechanical ventilation and 90.5% (95% CI, 31.5-99.6) for death. The bias-indicators showed increased risk during the period 0-13 days after single dose (**Table 2**).

**Table 2.** Adjusted vaccine effectiveness of Ad26.COV2.S against symptomatic COVID-19 and clinical outcomes.

|  | **Adjusted OR (95 CI)** | **Adjusted VE (95% CI)** |
| --- | --- | --- |
| **Symptomatic COVID19 (n=11,817)** |  |  |
| Unvaccinated |  |  |
| 0-13 days after single dose | 1.56 (1.27 to 1.91) | -55.8% (-90.8 to -27.3) |
| 14-27 days after single dose | 0.73 (0.57 to 0.91) | 27.4% (8.7 to 42.7) |
| ≥28 days after single dose | 0.49 (0.37 to 0.64) | 50.9% (35.5-63.0) |
| **Hospital admission (8,067)** |  |  |
| Unvaccinated |  |  |
| 0-13 days after single dose | 1.37 (0.78 to 2.29) | -37.3% (-129.1 to 22.1) |
| 14-27 days after single dose | 0.67 (0.30 to 1.29) | 33.5% (-29.1 to 69.8) |
| ≥28 days after single dose | 0.27 (0.09 to 0.65) | 72.9% (35.1 to 91.1) |
| **ICU admission (n=7,745)** |  |  |
| Unvaccinated |  |  |
| 0-13 days after single dose | 1.08 (0.34 to 2.71) | -7.5% (-170.6 to 66.1) |
| 14-27 days after single dose | 0.44 (0.07 to 1.53) | 56.0% (-52.8 to 93.1) |
| ≥28 days after single dose | 0.08 (0 to 0.45) | 92.5% (54.9 to 99.6) |
| **Use of invasive mechanical ventilation (n=7,697)** |  |  |
| Unvaccinated |  |  |
| 0-13 days after single dose | 1.1 (0.28 to 3.23) | -10.3% (-222.9 to 72.2) |
| 14-27 days after single dose | 0.35 (0.02 to 1.75) | 65.2% (-74.7 to 98.1) |
| ≥28 days after single dose | 0.11 (0 to 0.82) | 88.7% (17.9 to 99.5) |
| **Death (n=7,728)** |  |  |
| Unvaccinated |  |  |
| 0-13 days after single dose | 1.71 (0.58 to 4.16) | -70.7% (-316.5 to 42.2) |
| 14-27 days after single dose | 0.51 (0.07 to 1.92) | 48.9% (-92.3 to 92.5) |
| ≥28 days after single dose | 0.09 (0 to 0.68) | 90.5% (31.5 to 99.6) |

VE - vaccine effectiveness

## Discussion

We observed an effectiveness of 51% after 28 days of a single dose of Ad26.COV2.S against symptomatic COVID-19 and higher protection against clinical outcomes. Our results should be interpreted with the following issues. We do not have sequencing of this sample, but Gamma was predominant and Delta was rapidly emerging during the study period in Brazil. We analyzed a short follow-up period, thus we cannot evaluate potential waning on effectiveness. The bias-indicator showed residual confounding, which can be explained by the extensive outbreak that was ongoing when the vaccination started, and could indicate that our effectiveness estimates are underestimated.^5^

Despite lacking precision on some estimates, a single dose of Ad26.COV2.S vaccine continues to protect specially for severe forms of COVID-19 in the context of new variants.

## Data Availability

Deidentified databases as well as the R codes will be deposited in the repository https://github.com/juliocroda/VebraCOVID-19

## Acknowledgment

We are grateful to the National Immunization Program, Ministry of Health, Brazil, for the vaccine supply to cover the mass campaign in the 13 municipalities in the internal borders of Mato-Grosso do Sul.

## Author Contributions

Dr Ranzani and Dr Croda had full access to all of the data in the study and take responsibility for the integrity of the data and the accuracy of the data analysis.

Concept and design: All authors.

Acquisition, analysis, or interpretation of data: All authors.

Drafting of the manuscript: Ranzani, Croda.

Critical revision of the manuscript for important intellectual content: All authors

Statistical analysis: Ranzani.

Obtained funding: NA.

Administrative, technical, or material support: Leite, Castilho, Resende.

Supervision: Croda.

## Additional Contributions

We thank Daniel Henrique Tsuha for automating database linkage and Victor Bertollo Gomes Porto for advice on study design.

